# A plasma proteomic signature for atherosclerotic cardiovascular disease risk prediction in the UK Biobank cohort

**DOI:** 10.1101/2024.09.13.24313652

**Authors:** Trisha P. Gupte, Zahra Azizi, Pik Fang Kho, Jiayan Zhou, Ming-Li Chen, Daniel J. Panyard, Rodrigo Guarischi-Sousa, Austin T. Hilliard, Disha Sharma, Kathleen Watson, Fahim Abbasi, Philip S. Tsao, Shoa L. Clarke, Themistocles L. Assimes

## Abstract

**Background:** While risk stratification for atherosclerotic cardiovascular disease (ASCVD) is essential for primary prevention, current clinical risk algorithms demonstrate variability and leave room for further improvement. The plasma proteome holds promise as a future diagnostic and prognostic tool that can accurately reflect complex human traits and disease processes. We assessed the ability of plasma proteins to predict ASCVD.

**Method:** Clinical, genetic, and high-throughput plasma proteomic data were analyzed for association with ASCVD in a cohort of 41,650 UK Biobank participants. Selected features for analysis included clinical variables such as a UK-based cardiovascular clinical risk score (QRISK3) and lipid levels, 36 polygenic risk scores (PRSs), and Olink protein expression data of 2,920 proteins. We used least absolute shrinkage and selection operator (LASSO) regression to select features and compared area under the curve (AUC) statistics between data types. Randomized LASSO regression with a stability selection algorithm identified a smaller set of more robustly associated proteins. The benefit of plasma proteins over standard clinical variables, the QRISK3 score, and PRSs was evaluated through the derivation of Δ AUC values. We also assessed the incremental gain in model performance using proteomic datasets with varying numbers of proteins. To identify potential causal proteins for ASCVD, we conducted a two-sample Mendelian randomization (MR) analysis.

**Result:** The mean age of our cohort was 56.0 years, 60.3% were female, and 9.8% developed incident ASCVD over a median follow-up of 6.9 years. A protein-only LASSO model selected 294 proteins and returned an AUC of 0.723 (95% CI 0.708-0.737). A clinical variable and PRS-only LASSO model selected 4 clinical variables and 20 PRSs and achieved an AUC of 0.726 (95% CI 0.712-0.741). The addition of the full proteomic dataset to clinical variables and PRSs resulted in a Δ AUC of 0.010 (95% CI 0.003-0.018). Fifteen proteins selected by a stability selection algorithm offered improvement in ASCVD prediction over the QRISK3 risk score [Δ AUC: 0.013 (95% CI 0.005-0.021)]. Filtered and clustered versions of the full proteomic dataset (consisting of 600-1,500 proteins) performed comparably to the full dataset for ASCVD prediction. Using MR, we identified 11 proteins as potentially causal for ASCVD.

**Conclusion:** A plasma proteomic signature performs well for incident ASCVD prediction but only modestly improves prediction over clinical and genetic factors. Further studies are warranted to better elucidate the clinical utility of this signature in predicting the risk of ASCVD over the standard practice of using the QRISK3 score.

## Introduction

Atherosclerotic cardiovascular disease (ASCVD) is one of the leading causes of morbidity and mortality worldwide. A substantial contributor to the high burden of disease includes suboptimal risk stratification coupled with the inefficient application of established primary prevention strategies^1-3^. While ASCVD outcomes have improved in recent decades, clinical risk algorithms have historically demonstrated variability and leave room for further improvement^4^. Among ASCVD clinical risk prediction models currently in use, the UK-based QRISK3 cardiovascular clinical risk score has notably demonstrated excellent performance in its validation cohort and in the UK Biobank (UKB)^5,6^. Unlike other models, the QRISK3 score incorporates significant past medical history and current medication use in addition to standard clinical variables.

In the last decade, high-throughput profiling of circulating plasma proteins has emerged as a powerful tool for both predicting and understanding the underlying biology of complex human traits^7^. By capturing dynamic changes in protein expression, proteomic profiling is well-advantaged to reflect interactions between individuals’ genetics, environment, lifestyle, and more. Further, the incorporation of proteomic profiling data to traditional cardiovascular risk factors has also been shown to enhance prediction of cardiometabolic disease^8-10^. However, the ability of proteins to augment existing clinical risk prediction models as robust as the QRISK3 score has yet to be tested.

We aimed to evaluate the predictive value of clinical, genetic, and proteomic factors for ASCVD in the UKB. We hypothesized that the incorporation of proteomic data would provide modest improvement in prediction accuracy beyond that offered by the QRISK3 score, other clinical variables, and polygenic risk scores (PRSs).

## Methods

### Study population

We analyzed a cohort of UKB participants with normalized protein expression (NPX) data and excluded individuals with prevalent ASCVD, history of statin use, or in whom the QRISK3 score could not be computed. The study design of the UKB has been previously described extensively^11^. At participants’ baseline visits, trained healthcare providers conducted verbal interviews and administered questionnaires to obtain information on past medical history, family history, lifestyle, and sociodemographic and psychosocial factors. Physical measures along with the collection of blood, urine, and saliva samples were also obtained. By integrating participants’ electronic health record (EHR) data, health outcomes data including outpatient and inpatient International Classification of Disease, Tenth Revision (ICD-10) codes are available within the database. The UKB received ethical approval from the Northwest Multicenter Research Ethics Committee and obtained informed consent from all participants at the time of recruitment.

### Measurement of protein biomarkers

The UKB conducted proteomic profiling in a random sampling of UKB participants with plasma samples collected at baseline visits. Using the antibody-based Proximity Extension Assay (PEA) by Olink, the UKB measured NPX data for a total of 2,923 proteins. The sample handling, processing, and quality control protocols implemented by the UKB have been previously described in a summary document (biobank.ndph.ox.ac.uk/ukb/ukb/docs/PPP_Phase_1_QC_dataset_companion_doc.pdf) and in two publications^12,13^. We identified GLIPR1, NPM1, and PCOLCE as proteins with a high degree of missingness (> 50%) and excluded these from analysis. The remaining missing NPX values were imputed with their mean values. All NPX values were provided as log-transformed by the UKB and standardized by us prior to analysis.

### Measurement of the outcome

In accordance with the QRISK3 score, we defined ASCVD as the composite outcome of either transient ischemic attack (TIA), ischemic stroke, or coronary heart disease. The incidence of ASCVD was recorded using UKB’s first occurrences data (UKB categories 2401-2417), which is organized by ICD-10 codes and was generated by mapping primary care data (UKB category 3000), hospital inpatient data (UKB category 2000), death register records (UKB fields 40001 and 40002), and self-reported medical condition codes (UKB field 20002). The ICD-10 codes used to record ASCVD can be found in the **Supplementary File**.

### Measurement of clinical variables

Our primary clinical variable was the QRISK3 score, which incorporates several established clinical predictors to provide an individual’s 10-year predicted risk of developing ASCVD^5^. We computed the score using the QRISK3 function in the QRISK3 R package. We further considered additional clinical predictors measured at baseline including hemoglobin A1c (HbA1c), lipoprotein (a) (Lp(a)), LDL cholesterol, triglyceride levels, prior alcohol use, and physical activity status. Lastly, we considered 36 standard polygenic risk scores (PRSs) calculated with genome wide data on DNA sequence variation, summarizing the genetic predisposition or liability to 36 health traits or disease conditions^14,15^. All clinical variables and PRSs were standardized prior to running analyses. A full list of variables included in our analyses can be found in the **Supplementary File**.

### Statistical analyses

A flowchart of the study design and analysis plan is shown in **Figure 1**. We used least absolute shrinkage and selection operator (LASSO) regression to evaluate the relative ability of clinical variables, PRSs, and proteins to associate with incident ASCVD. We randomly divided the cohort into a training set (70%) and test set (30%) before building five LASSO models using 10-fold cross validation. These five LASSO models included a model with clinical variables alone, a model with PRSs alone, a model with proteins alone, a combined model with clinical variables and PRSs, and a combined model in which proteins were added to clinical variables and PRSs. We compared area under the curve (AUC) statistics to assess the relative ability of each model to predict ASCVD. To ascertain the incremental value offered by proteins beyond clinical variables and PRSs, we calculated Δ AUC values and generated a corresponding 95% confidence interval by bootstrapping 1,000 samples.

**Figure 1.**
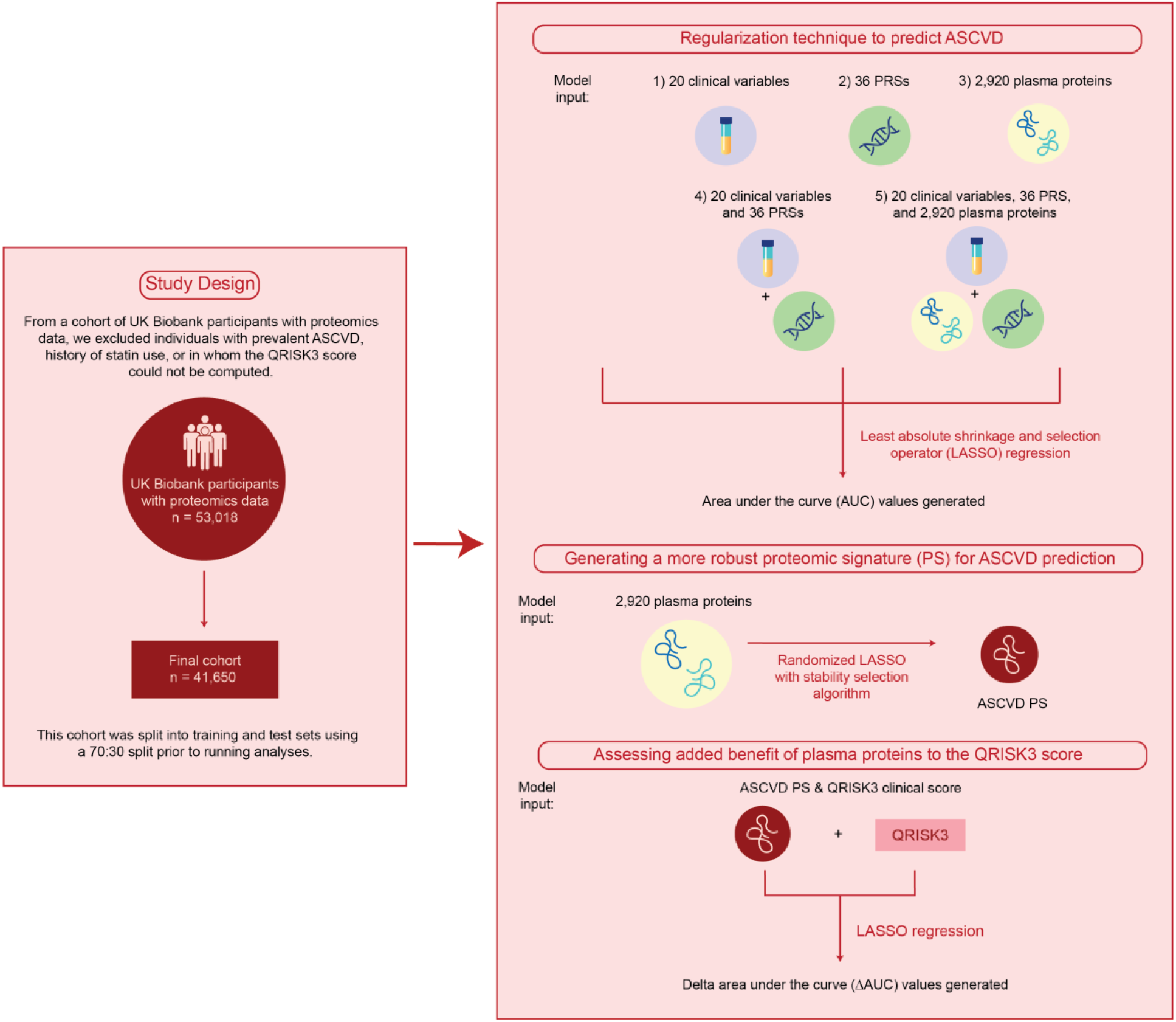
Study design and analysis workflow. **Abbreviations:** ASCVD: atherosclerotic cardiovascular disease, PRSs: polygenic risk scores

We next utilized the R package stabs to run the randomized LASSO stability selection (RLSS) algorithm, which was initially presented by Meinhausen and Bühlmann and later refined by Shah and Samsworth^16,17^. This algorithm was applied in the training set to develop a more robust proteomic signature (PS) for ASCVD prediction. We used default parameters when applying this algorithm, which included a weakness value of 0.8, a cutoff value of 0.8, and a per-family error rate of two. To assess the predictive value of this PS, we calculated the AUC of a LASSO model incorporating this smaller set of proteins in the test set. We subsequently calculated a Δ AUC value with a 95% confidence interval to determine the added predictive benefit of this PS in ASCVD prediction beyond that offered by QRISK3.

To further explore correlation structure and the incremental improvement in model performance with varying sizes of proteomic datasets, we used two methods to reduce the number of proteomic predictors. First, we implemented a filtering-based approach and computed a correlation matrix of all 2,920 proteins to identify pairs of proteins with a correlation value > 0.3 and > 0.5. We randomly removed one protein from each pair to form two smaller datasets of about 600 and 1,500 proteins, respectively. Second, we used a clustering-based approach and applied principal component analysis (PCA) and K-means clustering to form 600 and 1,500 clusters of proteins. From each cluster, we randomly selected a protein to form two additional smaller datasets of 600 and 1,500 proteins, respectively. Through standard LASSO regression, we evaluated ASCVD prediction performance of all four proteomic datasets by generating AUCs.

Finally, we used two-sample Mendelian randomization (MR) analysis to identify potential causal proteins for ASCVD. In a cohort of 15,016 UKB participants, we performed genome-wide association studies (GWASs) for all 2,920 proteins. Effect estimates for ASCVD were obtained by performing a GWAS analysis of ASCVD (defined as either a TIA, ischemic stroke, or coronary heart disease) in a cohort of UKB participants of European ancestry who did not have proteomics data available (n_cases_ = 64,386 & n_controls_ = 299,887). A full description of the methods used for both GWAS analyses can be found in the **Supplementary File**.

The inverse variance weighted method (IVW) served as our primary approach for the MR analysis. In any MR analysis, there are three assumptions which should be satisfied: 1) the genetic variants used as instrumental variables should be associated with the outcome of interest, 2) the genetic instruments should not be associated with other confounder variables, and 3) the genetic instruments should affect the outcome only via the exposure of interest rather than through alternative pathways. While MR analyses utilizing *cis*-genetic instruments have typically been found to satisfy these assumptions, we conducted additional analyses to address these assumptions^18^. To address the first assumption, we used established methods to calculate the proportion of variance explained and *F* statistic (with equations provided in the **Supplementary File**). Various sensitivity analyses were also conducted, including the MR-Egger method, which we used to calculate the MR-Egger intercept and assess for pleiotropy. All analyses were conducted using the TwoSampleMR package in R.

## Results

### Cohort characteristics

We analyzed NPX data of 2,920 proteins in a total of 41,650 participants. Baseline characteristics for the cohort are shown in **Table 1**. The mean age at recruitment was 56.0 years (SD, 8.2 years), 60.3% were female, and 93.1% were of self-reported white ethnicity. Over a median follow-up of 6.9 years, 9.8% developed incident ASCVD.

**Table 1.**
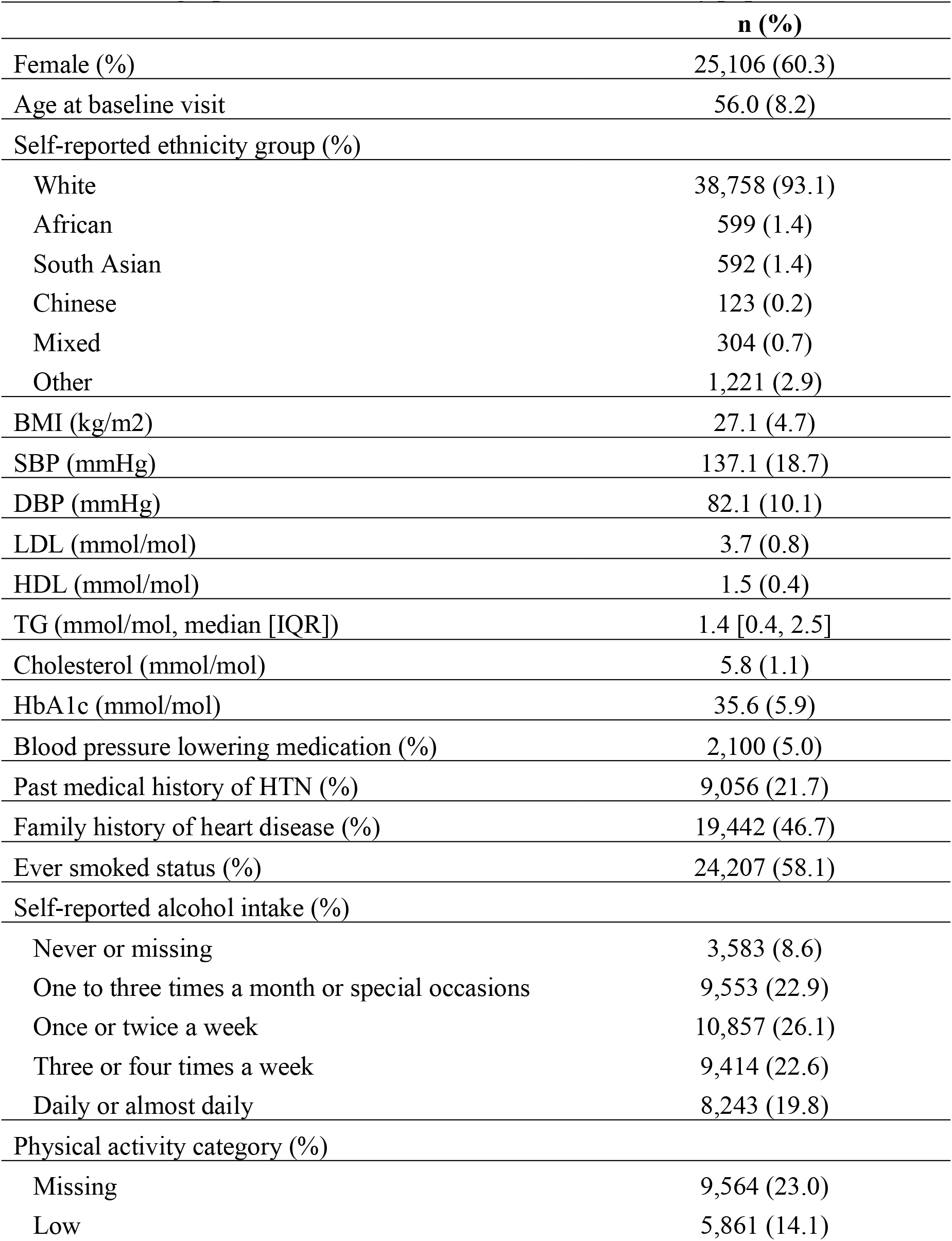
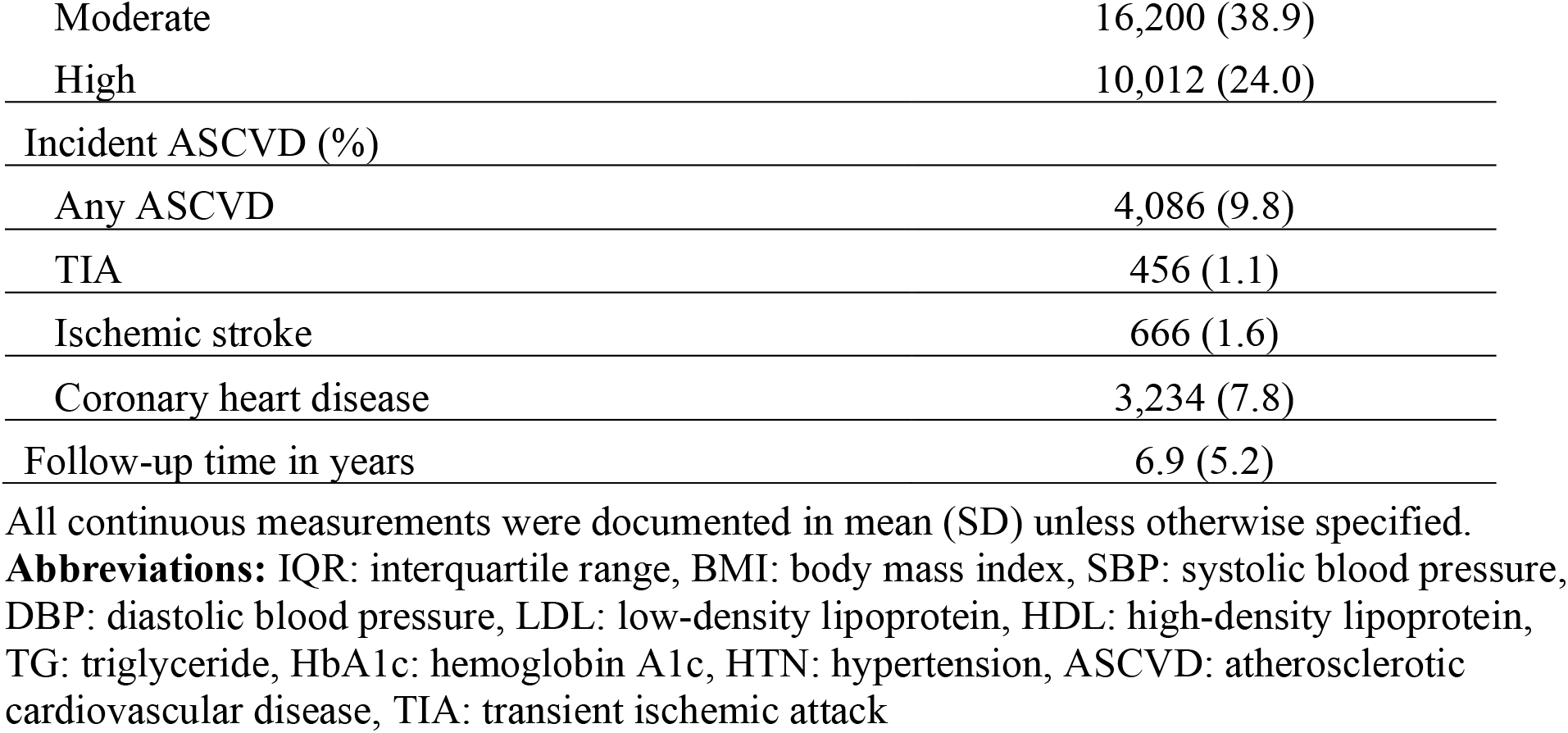
Demographics and clinical characteristics of the study population.

|  | **n (%)** |
| --- | --- |
| Female (%) | 25,106 (60.3) |
| Age at baseline visit | 56.0 (8.2) |
| Self-reported ethnicity group (%) |  |
| White | 38,758 (93.1) |
| African | 599 (1.4) |
| South Asian | 592 (1.4) |
| Chinese | 123 (0.2) |
| Mixed | 304 (0.7) |
| Other | 1,221 (2.9) |
| BMI (kg/m<sup>2</sup>) | 27.1 (4.7) |
| SBP (mmHg) | 137.1 (18.7) |
| DBP (mmHg) | 82.1 (10.1) |
| LDL (mmol/mol) | 3.7 (0.8) |
| HDL (mmol/mol) | 1.5 (0.4) |
| TG (mmol/mol, median [IQR]) | 1.4 [0.4, 2.5] |
| Cholesterol (mmol/mol) | 5.8 (1.1) |
| HbA1c (mmol/mol) | 35.6 (5.9) |
| Blood pressure lowering medication (%) | 2,100 (5.0) |
| Past medical history of HTN (%) | 9,056 (21.7) |
| Family history of heart disease (%) | 19,442 (46.7) |
| Ever smoked status (%) | 24,207 (58.1) |
| Self-reported alcohol intake (%) |  |
| Never or missing | 3,583 (8.6) |
| One to three times a month or special occasions | 9,553 (22.9) |
| Once or twice a week | 10,857 (26.1) |
| Three or four times a week | 9,414 (22.6) |
| Daily or almost daily | 8,243 (19.8) |
| Physical activity category (%) |  |
| Missing | 9,564 (23.0) |
| Low | 5,861 (14.1) |
| Moderate | 16,200 (38.9) |
| High | 10,012 (24.0) |
| Incident ASCVD (%) |  |
| Any ASCVD | 4,086 (9.8) |
| TIA | 456 (1.1) |
| Ischemic stroke | 666 (1.6) |
| Coronary heart disease | 3,234 (7.8) |
| Follow-up time in years | 6.9 (5.2) |

All continuous measurements were documented in mean (SD) unless otherwise specified.
**Abbreviations:** IQR: interquartile range, BMI: body mass index, SBP: systolic blood pressure, DBP: diastolic blood pressure, LDL: low-density lipoprotein, HDL: high-density lipoprotein, TG: triglyceride, HbA1c: hemoglobin A1c, HTN: hypertension, ASCVD: atherosclerotic cardiovascular disease, TIA: transient ischemic attack

### Standard LASSO regression

Consistent with prior reports, the QRISK3 score performed very well in predicting ASCVD with an AUC of 0.720 (95% CI 0.706-0.734) (**Fig. 2**). When additional clinical variables were added to QRISK3, the AUC marginally increased at 0.724 (95% CI 0.710-0.738). A LASSO model built on PRSs alone did not perform well with an AUC of 0.575 (95% CI 0.558-0.591) while a LASSO model incorporating the full proteomic dataset on its own performed comparably to the clinical variable-only model with an AUC of 0.723 (95% CI 0.717-0.745). In our combined models, we observed that the addition of PRSs to clinical variables resulted in a slightly higher AUC point estimate of 0.731 (95% CI 0.717-0.745). Incorporating the full proteomic dataset in addition to clinical variables and PRSs modestly improved the AUC to 0.741 (95% CI 0.727-0.755) and resulted in a Δ AUC of 0.010 (95% CI 0.003-0.018). A full list of the clinical variables, PRSs, and proteins selected by these LASSO models can be found in **Supp. Table 1**.

**Figure 2.**
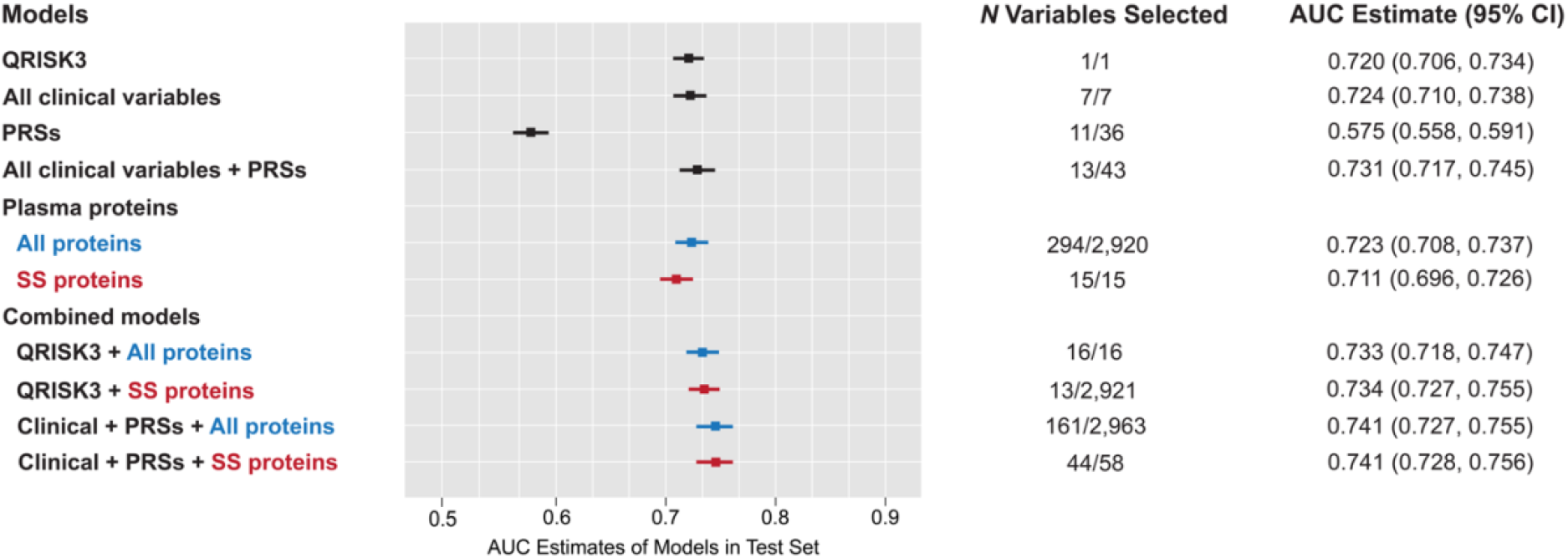
Area under the curves (AUCs) of clinical variables, polygenic risk scores, and plasma proteins. **Footnote:** Models performed using training and test sets in the study cohort. “All proteins” in blue represent the full proteomic dataset. “SS proteins” in red refer to proteins selected by a randomized LASSO regression model with stability selection algorithm. **Abbreviations:** LASSO: least absolute shrinkage and selection operator, AUC: area under the curve, PRSs: polygenic risk scores, SS: stability selection

### Stability selection with randomized LASSO regression

A randomized LASSO stability selection (RLSS) analysis selected 15 proteins, which spanned a wide range of known functions (**Fig. 3**). These stability selection proteins offered most of the improvement in ASCVD prediction explained by the full proteomic dataset with an AUC of 0.711 (95% CI 0.696-0.726). This robust proteomic signature also offered modest improvement in ASCVD prediction over that provided by the QRISK3 score [Δ AUC: 0.013 (95% CI 0.005-0.021)].

**Figure 3.**
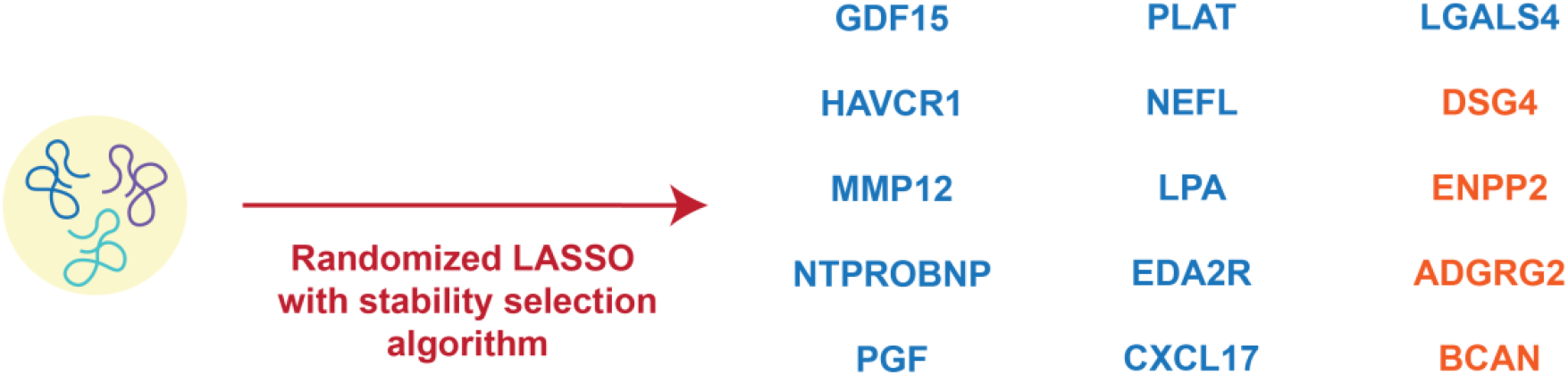
Plasma proteins identified by a stability selection algorithm with randomized LASSO regression. **Footnote:** Proteins listed in blue were positively associated with atherosclerotic cardiovascular disease and those listed in orange were negatively associated. **Abbreviations:** LASSO: least absolute shrinkage and selection operator

### Clustering and filtering analyses

We assessed and compared the prediction performance of smaller proteomic datasets formed through filtering-based or clustering-based approaches to that of the full proteomic dataset provided by the UKB [AUC of 0.723 (95% CI 0.708-0.737)] (**Supp. Fig. 1**). A smaller dataset (consisting of 600 proteins) formed through a filtering-based approach with a correlation threshold of > 0.3 performed the worst with an AUC of 0.691 (95% CI 0.675-0.706). Despite consisting of approximately the same number of proteins (645 vs. 600), a smaller dataset formed through a clustering-based approach with the creation of 600 clusters performed better with an AUC of 0.705 (95% CI 0.690-0.720). Finally, datasets formed through a filtering-based approach with a correlation threshold of > 0.5 and through a clustering-based approach with the creation of 1,500 clusters performed comparably to the full proteomic dataset with AUCs of 0.719 (95% CI 0.704-0.734) and 0.717 (95% CI 0.702-0.732), respectively.

### Two-sample Mendelian randomization analysis

To identify potentially causal proteins for ASCVD, we conducted a two-sample Mendelian randomization (MR) analysis. Of 2,920 proteins, we identified genome-wide significant *cis*-protein quantitative loci (*cis*-pQTLs) for 1,745 based on a significance threshold of 5 x 10^-8^. The minimum *F* statistic of our genetic instruments was 27.7 (**Supp. Table 2**). Effect estimates for ASCVD were obtained by running a GWAS in 364,273 UKB participants of European ancestry using REGENIE (**Supp. Fig. 2a-b**). We identified 11 proteins as having a potentially causal effect on ASCVD based on a Bonferroni-corrected threshold (p-value = 4.31 x 10^-5^) (**Fig. 4**). For several proteins in which the initial number of associated SNPs was low (nSNPs < 3), we were not able to obtain results for sensitivity analyses. For proteins with a higher number of initial associated SNPs, however, we found that results from the IVW method generally aligned with results from other sensitivity analyses (**Supp. Fig. 4)**. Full results for the two-sample MR analysis including annotations of whether each protein tested was selected by a standard LASSO model or the RLSS algorithm can be found in **Supp. Table 2**.

**Figure 4.**
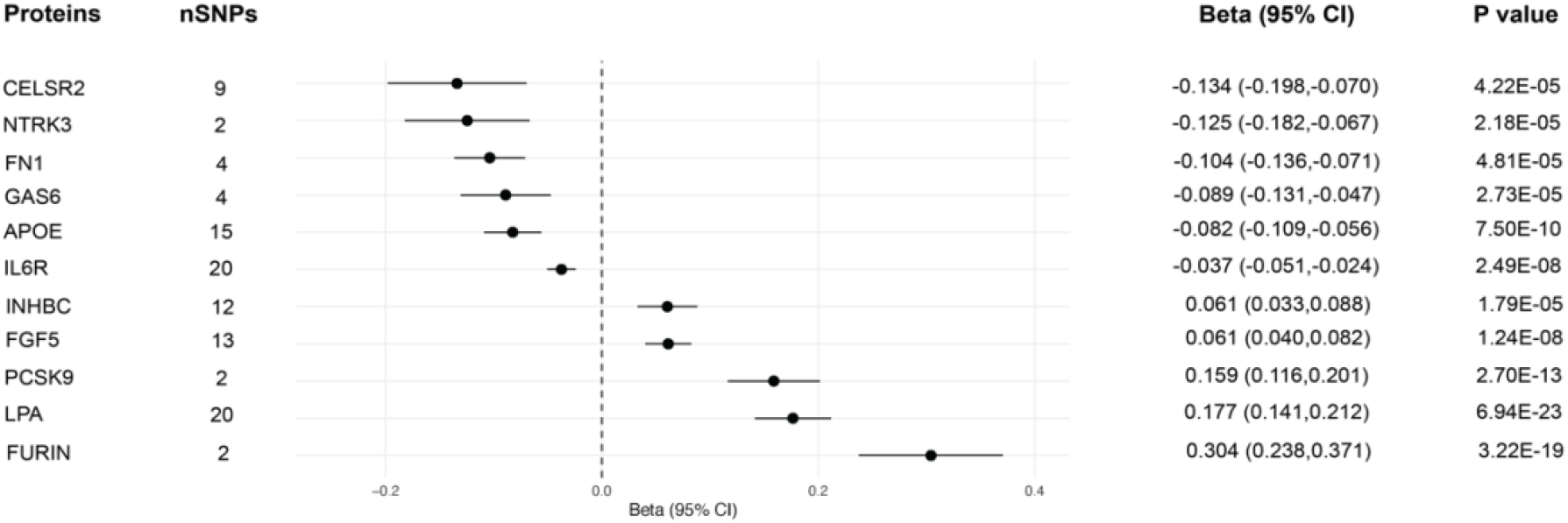
Potentially causal proteins for atherosclerotic cardiovascular disease identified in a two-sample Mendelian randomization analysis. **Footnote:** Forest plot of potentially causal proteins for atherosclerotic cardiovascular disease based on a Bonferroni-corrected threshold (p-value = 4.31 x 10^-5^). **Abbreviations:** SNP: single nucleotide polymorphism, 95% CI: 95% confidence interval

## Discussion

We evaluated the relative ability of clinical variables, PRSs, and proteins to predict ASCVD by utilizing high-throughput proteomic profiling data provided by the UKB. In this study, we investigated whether the incorporation of proteomic data enhanced prediction offered by an existing clinical risk prediction model, as well as other clinical and genetic factors. We highlight three primary sets of findings from our results.

First, we found that a protein-only model derived by standard LASSO regression performed comparably to the QRISK3 score, which performed exceptionally well on its own. When additional clinical variables and PRSs were combined with QRISK3, ASCVD prediction was comparable to performance offered by QRISK3 alone. The further addition of proteins to clinical variables and PRSs resulted in a modest improvement in prediction. Unlike most ASCVD risk prediction models, the UK-based QRISK3 cardiovascular clinical risk score captures a significant portion of an individual’s past medical history and current medication use, which explains its strong performance in UK-based populations such as its validation cohort and in the UKB^5,6^. In the setting of our baseline model already exhibiting strong performance, our observation of only modest improvements in ASCVD prediction with the addition of other clinical variables and multi-omic datasets is not surprising^19^. However, when practitioners may not have access to all the clinical information needed to compute a score generated by QRISK3, the comparable performance of our protein-only model suggests that proteins could potentially serve as an alternative tool for risk stratification purposes in the future.

Second, we show that a substantially smaller set of proteins selected by a stability selection algorithm accounted for most of the prediction performance offered by the full dataset. In addition to aiding in feature reduction, this algorithm also identifies more “stable” features that may be more transportable to other populations. Recently, others created a custom quantitative PEA panel measuring up to 21 proteins called the CVD-21 tool^20^. With this tool in mind, a future where a robust proteomic signature such as the one we describe is generated for risk prediction through absolute quantification is now foreseeable. Third, smaller proteomic datasets, created by filtering and clustering methods to reduce high degrees of correlation in the full dataset, do not meaningfully change the performance of ASCVD prediction. When looking to future implementations of plasma proteomic profiling, our findings suggest that the additional costs associated with measuring more than ∼1,500 proteins may be avoided without drastically affecting prediction performance.

Several proteins were repeatedly selected by standard LASSO models incorporating proteins alone, as well as proteins in addition to clinical variables and PRSs. In particular, GDF15, or growth/differentiation factor 15, has previously been associated with a host of diseases within the cardiometabolic spectrum^21,22^. With known functions in the suppression of food intake and inflammation, GDF15 is now an appealing drug target in the management of obesity, T2DM, and CVD^23,24^. LTBP2 also carried a large beta coefficient in our protein-only LASSO model. While this protein has not previously been associated with ASCVD, others have demonstrated its potential as a novel biomarker of heart failure and other diseases^25,26^. Interestingly, we also identified dementia-related proteins such as BCAN and NEFL as predictive of ASCVD as well^7,27^.

In addition to risk prediction, high-throughput proteomic profiling can further our understanding of the underlying biology of disease and aid in identifying novel drug targets. From our two-sample MR analysis, we corroborated several previously known causal targets of ASCVD including PCSK9, LPA, and IL6R^28-31^. Additionally, we identified FN1 as potentially causal, which has been demonstrated to enhance endothelial inflammation in atherosclerotic plaques^32^. Lastly, we identified CELSR2 as potentially causal for ASCVD. With known functions in serum lipid and cholesterol metabolism, variants in the *CELSR2* gene and its neighboring genes, *PSCR1* and *SORT1* were first identified in GWASs of cardiovascular disease over fifteen years ago^33^.

The well-documented reproducibility and stability of the Olink platform is a key strength of our study^34^. By selecting QRISK3, a robust cardiovascular clinical risk score, as our baseline model, we also contribute valuable insights on whether proteins can augment existing prediction models for ASCVD. Our study has some notable limitations including the lack of genetic diversity within the UKB. Our two-sample MR analysis was restricted to participants of European ancestry only. To avoid the further perpetuation of health disparities in biomedical research, future studies in more diverse populations are needed. Additionally, while previous studies have also highlighted the potential of plasma proteins to enhance cardiometabolic health prediction, we acknowledge potential limitations in relying on protein measurements in plasma rather than using protein measurements from human tissue^9^ .

In summary, our findings suggest that plasma proteomic profiling modestly enhances prediction of ASCVD beyond an already well-performing cardiovascular clinical risk score, QRISK3, as well as other routinely available clinical variables and PRSs. Further investigations in more diverse study populations are needed to better understand the potential benefits multi-omics data could provide for ASCVD prediction. We also show that utilizing more robust proteomic datasets does not appreciably affect prediction performance when compared to the full proteomic dataset provided by the UKB. Finally, we contribute a list of potentially causal proteins for ASCVD using expanded plasma proteomic from the UKB.

## Data Availability

All data produced in the present study are available upon reasonable request to the authors.

## Protein and Gene Abbreviations

GDF15: Growth/differentiation factor 15
LTBP2: Latent-transforming growth factor beta-binding protein 2
BCAN: Brevican core protein
NEFL: Neurofilament light polypeptide
PCSK9: Proprotein convertase subtilisin/kexin type 9
LPA: Apolipoprotein(a)
IL6R: Interleukin-6 receptor subunit alpha
FN1: Fibronectin
CELSR2: Cadherin EGF LAG seven-pass G-type receptor 2
*PSCR1*: Proline/serine-rich coiled-coil 1
*SORT1*: Sortilin

## Acknowledgement

The UKB received ethical approval from the Northwest Multicenter Research Ethics Committee and obtained informed consent from all participants at the time of recruitment. This study was conducted under UK Biobank application number 52374.

## Funding and Assistance

This study was supported by a grant from the National Institutes of Health 1R01DK114183. TPG was supported by the Sarnoff Cardiovascular Research Foundation Fellowship.

## Author Contributions

TPG and TLA conceived and designed the study. TPG, ZA, PFK, JZ, and carried out the analyses. TPG and TLA drafted the manuscript. TPG, ZA, PFK, JZ, MLC, DJP, RGS, ATH, DS, KW, FA, PST, SLC, and TLA verified the underlying data. TLA is responsible for the integrity of the work as a whole. All authors acquired and interpreted the data, critically revised the paper and had final responsibility for the decision to submit for publication.

