## Supplementary File for "A plasma proteomic signature for atherosclerotic cardiovascular disease risk prediction in the UK Biobank cohort"

### Supplementary Methods

*Study design and population*

Prevalent atherosclerotic cardiovascular disease (ASCVD) was defined as the composite outcome of either transient ischemic attack (TIA), ischemic stroke, or coronary heart disease at or before the baseline visit. Incident ASCVD was defined as having any of these conditions after the baseline visit. We used the following ICD-10 codes within UK Biobank's First Occurrences data to record the prevalence or incidence of these conditions.

| Condition | ICD-10 code |
|---|---|
| Transient ischemic attack | G45 |
| Ischemic stroke | I63 |
| Coronary heart disease | I21.0 – I21.9, I22.0-I22.9, I23.1-I23.6, I24.0-I24.9, I25.1-I25.9 |

*Measurement of clinical variables and polygenic risk scores*

Clinical variables used to compute the QRISK3 along with additional clinical variables included in least absolute selection and shrinkage (LASSO) regression models are listed below. Of note, the QRISK3 score uses family history of a heart attack in a first degree relative. As this information is not currently available within the UK Biobank's database, we included family history of any heart disease instead.

| Variables | UKB field code |
|---|---|
| *Clinical variables used for the QRISK3 clinical score* | |
| Sex | 31 |
| Age at recruitment | 21022 |
| Self-reported ethnicity | 21000 |
| Weight | 21002 |
| Height | 12144 |
| Smoking status | 20160, 3456 |
| Townsend deprivation index | 22189 |
| Systolic blood pressure | 4080 |
| Total cholesterol to HDL ratio | 30690, 30760 |
| Family history of heart disease | 20107, 20110, 20111 |
| Past medical history of T1DM | 30870 |
| Past medical history of T2DM | 30750 |
| Past medical history of erectile dysfunction | 132088 |
| Past medical history of migraine | 131052 |
| Past medical history of rheumatoid arthritis | 131848 |

| | |
|---|---|
| Past medical history of hypertension | 131286 |
| Past medical history of chronic kidney disease | 132032 |
| Past medical history of severe mental illness | 130874, 130892 |
| Past medical history of systemic lupus erythematosus | 131894 |
| Past medical history of atrial fibrillation | 131350 |
| Use of blood pressure-lowering medications | 20003 |
| Use of atypical antipsychotics | 20003 |
| Use of steroid medications | 20003 |
| *Additional clinical variables used* | |
| Glycated hemoglobin | 30750 |
| Self-reported alcohol use | 1558 |
| Physical activity category | 22040 |
| Triglycerides | 30870 |
| LDL cholesterol | 30780 |
| Lp(a) | 30790 |
| *Standard polygenic risk scores (PRS) used in all three subcohorts* | |
| Standard PRS for age at menopause | 26202 |
| Standard PRS for age-related macular degeneration | 26204 |
| Standard PRS for age at Alzheimer's disease | 26206 |
| Standard PRS for asthma | 26210 |
| Standard PRS for atrial fibrillation | 26212 |
| Standard PRS for bipolar disorder | 26214 |
| Standard PRS for body mass index | 26216 |
| Standard PRS for bowel cancer | 26218 |
| Standard PRS for breast cancer | 26220 |
| Standard PRS for cardiovascular disease | 26223 |
| Standard PRS for celiac disease | 26225 |
| Standard PRS for coronary artery disease | 26227 |
| Standard PRS for Chron's disease | 26229 |
| Standard PRS for epithelial ovarian cancer | 26232 |
| Standard PRS for estimated bone mineral density t-score | 26234 |
| Standard PRS for glycated hemoglobin | 26238 |
| Standard PRS for height | 26240 |
| Standard PRS for high-density lipoprotein cholesterol | 26242 |
| Standard PRS for hypertension | 26244 |
| Standard PRS for intraocular pressure | 26246 |
| Standard PRS for ischemic stroke | 26248 |
| Standard PRS for low-density lipoprotein cholesterol | 26250 |
| Standard PRS for melanoma | 26252 |

| | |
|---|---|
| Standard PRS for multiple sclerosis | 26254 |
| Standard PRS for osteoporosis | 26258 |
| Standard PRS for Parkinson's disease | 26260 |
| Standard PRS for primary open angle glaucoma | 26265 |
| Standard PRS for prostate cancer | 26267 |
| Standard PRS for psoriasis | 26269 |
| Standard PRS for resting heart rate | 21150 |
| Standard PRS for rheumatoid arthritis | 26273 |
| Standard PRS for schizophrenia | 26275 |
| Standard PRS for systemic lupus erythematosus | 26278 |
| Standard PRS for total cholesterol | 21151 |
| Standard PRS for total triglyceride | 21152 |
| Standard PRS for type 1 diabetes | 26283 |
| Standard PRS for type 2 diabetes | 26285 |
| Standard PRS for ulcerative colitis | 26287 |
| Standard PRS for venous thromboembolic disease | 26289 |

Footnote:
a: For self-reported ethnicity, individuals who identified as British, Irish, or any other White background were classified as White or not stated; those who identified as African or any other Black background were classified as Black African; those who identified as Indian were classified as Indian; those who identified as Pakistani were classified as Pakistani; those who identified as Bangladeshi were classified as Bangladeshi; those who identified as Asian or Asian British were identified as Other Asian; those who identified as having a mixed background were classified as Mixed; all other individuals were classified as Other ethnic group. b: For both systolic blood pressure, two measurements were taken through an automated reading a few moments part. These measurements were averaged together. We also included the standard deviation of these measurements as a variable for the QRISK3 score. c: Self-reported medication data was used to classify the use of several medications. Statin medication use was recorded if atorvastatin, rosuvastatin, simvastatin, fluvastatin, pravastatin, velastatin, or eptastatin werelisted. Corticosteroid medication use was recorded if prednisolone, betamethasone, cortisone, depo-medrone, dexamethasone, deflazacort, efcortesol, hydrocortisone, methylprednisolone, or triamcinolone was listed. Lastly, second-generation antipsychotic medication was recorded if amisulpride, aripiprazole, clozapine, lurasidone, olanzapine, paliperidone, quetiapine, risperidone, sertindole, or zotepine was listed. d: Physical activity was defined as the summed Metabolic Equivalent Task (MET) minutes per week score. Individuals with a MET score of >3,000 minutes per week were classified as having high physical activity, those with a MET score between 600-3,000 minutes per week were classified as having moderate physical activity, and those with a MET score of <600 minutes per week were classified as having low physical activity. e: The 36 standard polygenic risk scores (PRSs) listed were generated by the UK Biobank using external genome-wide association study data.

*Statistical Analyses*

*Two-sample Mendelian randomization analysis: Cis-protein quantitative trait loci analysis*

We performed genome-wide association studies (GWASs) for all 2,920 proteins in a cohort of 15,016 UK Biobank participants to identify *cis*-protein quantitative trait loci (cis-pQTLs). Prior to GWAS analysis, SNPs with a minor allele frequency (MAF) < 0.001 or an imputation quality score < 0.3 were filtered. Individual NPX values were inverse-rank normalized prior to GWAS analysis. The GWAS analyses were performed using the REGENIE software, adjusting for several covariates: age, $age^2$, sex, proteomic batch, UKB recruitment center, UKB genotype array, the time interval between blood sampling and measurement, and the first 20 genetic principal components[1]. *Cis*-protein quantitative trait loci (*cis*-pQTLs) were defined as SNPs located within 1 Mb on either side of the protein-coding gene. We pruned SNPs based on linkage disequilibrium with a $r^2$ threshold of 0.01. We also adopted a significance threshold of $5 \times 10^{-8}$ for defining genetic instruments.

*Two-sample Mendelian randomization analysis: Obtaining effect estimates for atherosclerotic cardiovascular disease*

To obtain effect estimates for ASCVD, we ran a GWAS analysis of ASCVD in a cohort of UK Biobank participants ($n_{cases}$ = 64,386 & $n_{controls}$ = 299,887). To avoid sample overlap with the group in which *cis*-pQTLs were identified in, we excluded participants who underwent proteomic profiling. The GWAS analysis was performed with the REGENIE software, and was adjusted for age, sex, genotype batch, and the first 10 genetic principal components[1]. We filtered SNPs with a MAF > 0.01, minor allele count (MAC) < 100, genotype missingness > 0.1, or a Hardy-Weinberg equilibrium p-value > $10^{-15}$. Samples with more than 10% missingness were also filtered for quality control.

*Two-sample Mendelian randomization analysis: Calculating proportion of variance explained and F-statistics*

We calculated the proportion of phenotypic variance ($q^2$), the total proportion of variance explained ($R^2$), and the *F* statistic using previously established methods described by others[2,3].

$$q^2 = 2 \times MAF \times (1 - MAF) \times \beta^2$$

where $q^2$ is the proportion of phenotypic variance, MAF is the minor allele frequency, and β is the effect size for a given SNP.

$$R^2 = (2 \times MAF \times (1-MAF) \times \beta^2) / q^2$$

where $R^2$ is the total proportion of variance explained, MAF is the minor allele frequency, $q^2$ is the proportion of phenotypic variance, and β is the effect size for a given SNP.

$$F = R^2 *(N-1-k) / ((1- R^2) \times k)$$

where F is the *F* statistic, $R^2$ is the total proportion of variance explained, N is the sample size, and k is the number of SNPs included in the instrument.

**Supplementary Figure 1. Area under the curves (AUCs) using filtered and clustered proteomic datasets**

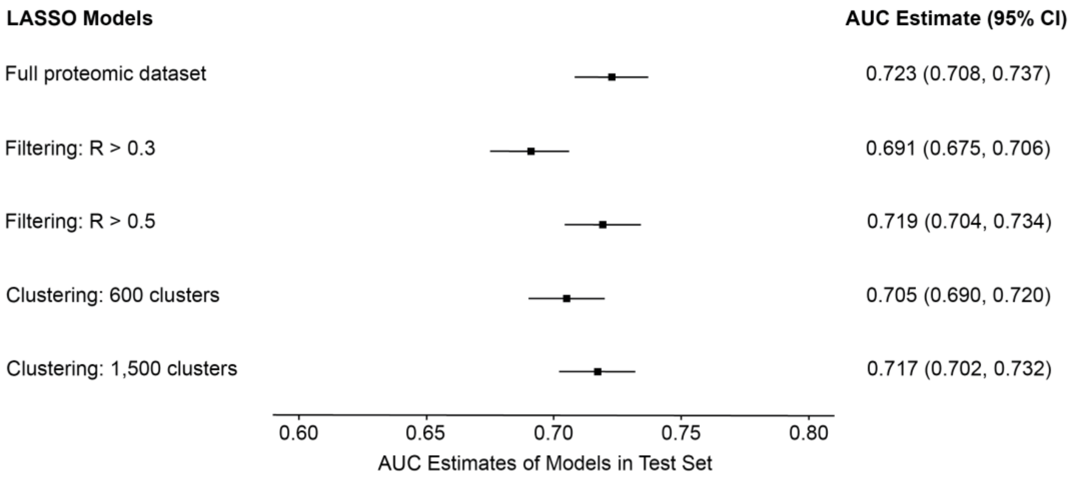


**Footnote:** Models performed in the training and test sets using filtered and clustered versions of the full proteomic dataset. **Abbreviations:** LASSO: least absolute shrinkage and selection operator, AUC: area under the curve, 95% CI: 95% confidence interval

**Supplementary Figure 2a-b. Manhattan and Q-Q plots for genome-wide association studies of atherosclerotic cardiovascular disease in UKB participants of European ancestry**

**a**

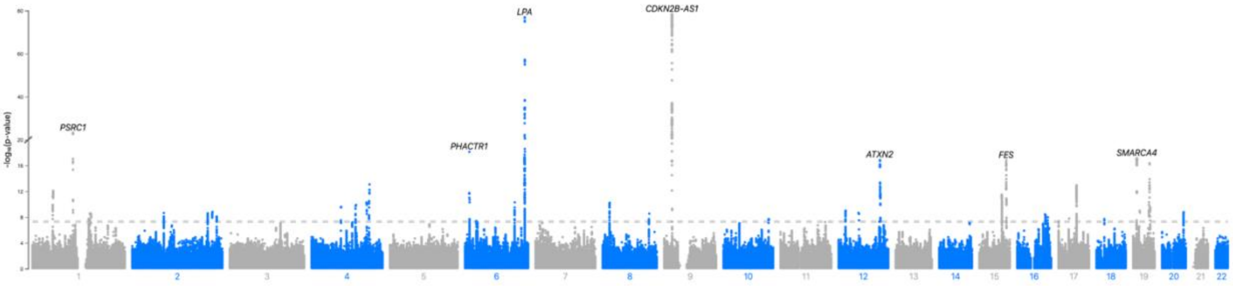


**b**

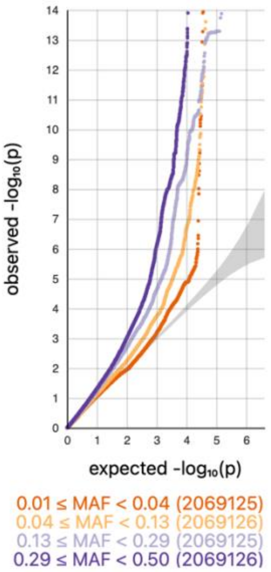


**Footnote:** (a) Manhattan plot of GWAS for atherosclerotic cardiovascular disease. (b) Q-Q plot of GWAS for atherosclerotic cardiovascular disease.

**Supplementary Figure 3. Cross-validation plots of LASSO regression models**

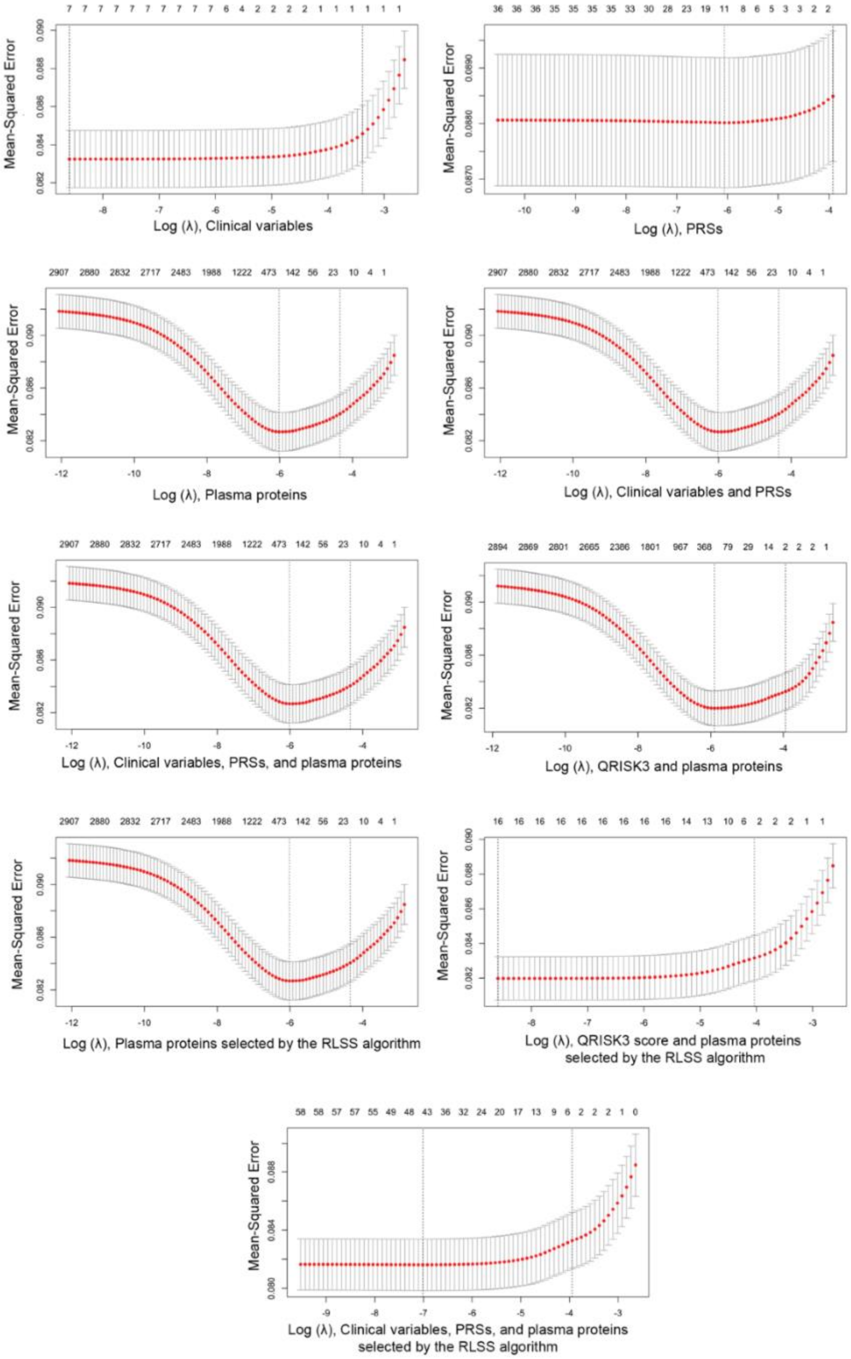


**Footnote:** Cross-validation plots of LASSO regression models in test data sets with atherosclerotic cardiovascular disease as the outcome of interest. Upper and lower standard deviation curves along the $\lambda$ sequence (error bars) are shown. **Abbreviations:** PRSs: polygenic risk scores

**Supplementary Figure 4. Potentially causal proteins for ASCVD**

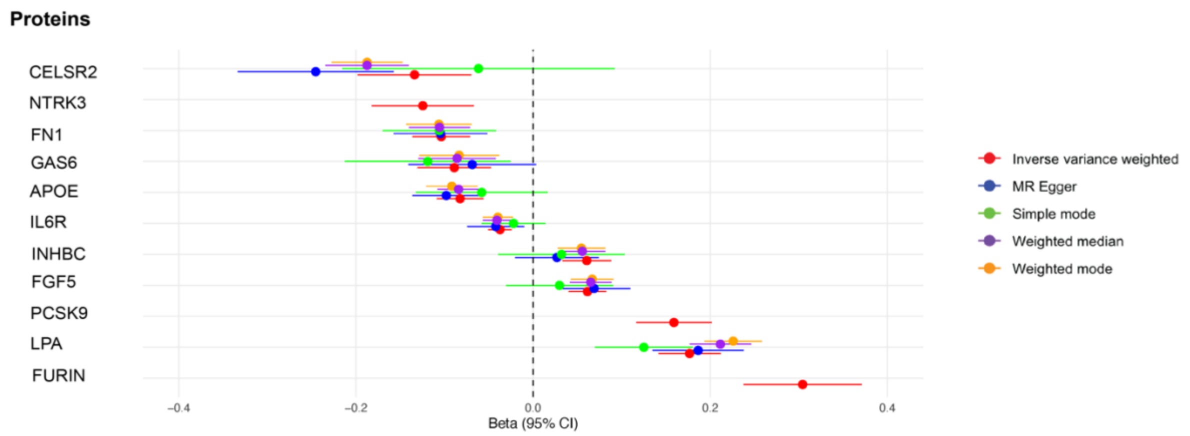


**Footnote:** Forest plot of potentially causal proteins for ASCVD based on Bonferroni-corrected threshold (p-value = $4.31 \times 10^{-5}$) using the inverse variance weighted method with results from other sensitivity analyses. Red denotes results from the inverse variance weighted method, blue from the MR-Egger method, green from the simple mode method, purple from the weighted median method, and orange from the weighted mode method. **Abbreviations:** 95% CI: 95% confidence interval
